# Tracking the COVID-19 vaccine equity, distribution, and cases in the global south

**DOI:** 10.1101/2022.12.19.22283681

**Authors:** Tigist Mekonnen Melesse, Thuy D. Nguyen, Getachew Mullu Kassa

## Abstract

The rapid development of vaccines against severe acute respiratory syndrome coronavirus 2 (SARS-CoV-2) has proved to make an important contribution in reducing both viral transmission and disease burden. In this study, we tracked the COVID-19 vaccine equity, distribution, and cases in global south countries using country-level data from Our World in Data using an event study analysis. We used data from 149 global south and 59 non-global south countries from January 2020 to May 2022. All non-global south and 90.32% of global south countries had universal availability of vaccines. The median time since the introduction of the first COVID-19 vaccine in the global south was almost eight weeks later than in non-global south countries. The median number of people fully vaccinated per hundred (68.8 vs 50.31), and the total number of boosters administered per hundred (45.7 vs. 13.02) were higher in non-global south countries compared to global south countries. Using the event study analysis, we found a significant reduction of COVID-19 new cases and deaths after the first COVID-19 vaccination rollout compared to the baseline in global south countries, average coefficient p-value <0.001. Programs aiming at improving vaccine access and distribution to global south countries are essential to effectively control COVID-19.

## 1. Introduction

The coronavirus pandemic (COVID-19) took over 6.5 million lives and over 630 million confirmed cases globally as of October 19, 2022 (WHO 2022). In 2020 only, 8.8 percent of global working hours were lost, which is equivalent to 255 million people losing their jobs (ILO Monitor 2020). By continent, employment was reduced by 8.3 percent in high-income countries, in upper-middle-income countries by 7.3 percent, in lower-middle-income countries by 11.3 percent, in low-income countries by 6.7 percent, in the United States of America by 4.7 percent, and in Africa by 4.1 percent (Gopinath 2020; Bundervoet et al. 2022). As a whole, the job losses during COVID-19 was 12.8 percent compared to the job losses during the global recession 2007-2009 (10 percent) or the financial crisis 1997-2001 (8.6 percent). The coronavirus pandemic crisis has been recorded as one of the worst recessions (-3 percent) since the great depression (-1 percent) economic downturn (Shibata 2020, Gopinath 2020). The global economic loss (GDP) in 2020 and 2021 was 9 trillion US dollars, which is greater than the economies of Japan and Germany (Gopinath 2020).

The rapid development of vaccines against severe acute respiratory syndrome coronavirus 2 (SARS-CoV-2) has proved to make an important contribution to reducing both viral transmission and disease burden (Gupta et al. 2021). However, concerns related to vaccine equity become the biggest obstacle to addressing this global pandemic and future pandemics. The Director-General of the World Health Organization shared that 1 in 4 people are vaccinated against COVID-19 in high-income countries (HICs) compared to 1 in 500 vaccinated in low-and-middle-income countries (LMICs) (Upadhyay et al. 2022). In September 2020, the WHO planned to initially allocate covid vaccine to countries considering the proportion to their population size. After countries received the covid vaccine doses for 20% of their population, their covid risk will be considered for the successive vaccine distribution (Herzog et al. 2021). Countries were also permitted to pursue bilateral contracts with vaccine manufacturers. The latter resulted in uncoordinated approach and competition of countries for covid vaccines in the market (Herzog et al. 2021). Furthermore, a study that has been published in The Lancet Infectious Disease finds that if no vaccines had been distributed, an estimated number of 20 million lives will be lost in the first year (Watson et al. 2022).

The inventions of novel coronavirus disease 2019 (COVID-19) vaccines have been quicker than previous vaccine inventions (Keni et al. 2020). Because of the complexity of vaccine development, it may take up to 10-15 years for vaccine development (Cohut 2020). However, the COVID-19 vaccine development took under a year mainly because of worldwide cooperation (Cohut 2020). Pfizer vaccine was the first vaccine that received emergency use authorization against COVID-19 from Food and Drug Administration (FDA) on December 11, 2020 (Cohut 2020). Furthermore, the World Health Organization (WHO) has approved and publicized ten COVID-19 vaccines, including Pfizer/BioNTech, Oxford (AstraZeneca), Janssen (Johnson & Johnson), Sinopharm (Beijing), Moderna, Sinovac, Covishield (Oxford/AstraZeneca formulation), Novavax, Covaxin, and COVOVAX (Novavax formulation) (WHO 2022a).

Vaccinations against COVID-19 have resulted in a reduction of COVID-19-related hospitalization and deaths, especially among older adults (Moghadas et al. 2021). A growing body of empirical evidence have also shown a significant waning effect of the vaccines against infections (and transmission) at 12-16 weeks, with both Delta and Omicron variants (Bardosh et al. 2022; Fabiani et al. 2022; Chemaitelly et al. 2021), including with third-dose shots. According to the (WHO 2022d) report, as of 19 August 2022, a total of 12,814,704,622 vaccine doses vaccine doses have been administered. Vaccination against COVID-19 and prior infections has resulted in the mitigation of disease outbreaks (Moghadas et al. 2021; Suthar et al. 2022). Although equitable access to COVID-19 vaccines is critical to contain the pandemic, the global percentage of persons fully vaccinated with the last dose of the primary series of the vaccine was 62.47%, highest in the Western Pacific WHO region (83.98%) and lowest in Africa (19.27%) (WHO 2022f).

Comparing the vaccine acceptance rate between low, middle, and high-income countries, the average vaccine acceptance rate in low-and middle-income countries (LMIC) was higher (80.3%) than that of the United States (64.6%), for instance, (Machingaidze and Wiysonge 2021). This study, therefore, assesses vaccine equity, distribution, and cases in the global south countries, and findings will inform governments, policymakers, and pharmaceutical companies on these issues. We applied an event regression and difference-in-difference estimation strategies where such techniques allow us to exploit variations in timing by geographical units. The COVID-19 vaccine confirmation date by the WHO is used as an information event/policy to assess the variation in timing by countries.

Our data also has the various dates when the vaccines are available across countries including first available, and first available to certain age groups/disaggregated by age. Having this various date in the data also helps to apply the event study regression and difference-in-difference estimation techniques, as well as when we use time and country fixed effects to control for certain factors.

## 2. Lesson learned from the previous pandemic

Since 1796, where the basis of vaccination begun for smallpox, the development of vaccines played a great role in controlling the prevalence and incidence of Vaccine Preventable Diseases (VPD) (Saint-Victor and Omer 2013). Elimination of poliomyelitis in America and eradication of smallpox worldwide is due to the development of vaccine and vaccination programs (Dubé et al. 2013). In 1918/19, influenza pandemic killed more people more than any other disease outbreak in history (Lemon et al. 2005). A Nobel Laureate Frank Macfarlane Burnet who discovered acquired immunological tolerance estimated the death toll by influenza pandemic between 50 to 100 million people (Frank MacFarlane Burnet et al.1972) and the first vaccines were approved in 1945 (Lina 2008). Since the first reported cases in 1981, the HIV/AIDS has killed approximately 36.3 million people worldwide. Since the outbreak of Ebola virus pandemic in 1976, over 15,000 people died because of the disease (Breman et al. 2016). Until now, there is no licensed vaccines to present HIV/AIDS and Ebola virus diseases. The outbreak of the coronavirus (COVID-19) pandemic in 2019 where over 6 million people has killed until 2022, took less than one year to develop the vaccines (Cohut 2020).

Despite taking over 10 years to develop vaccines during the previous pandemic, the contribution of science (inventing vaccines) to eradicate smallpox in the 1970s and polio in the 1990s and the governments’ effective approach to maximize vaccine would be a good experience for the current and future pandemic. Give the COVID-19 vaccines were in use in less than one year of the outbreaks the virus, the low uptake rate of the vaccines in low-and-middle income countries (14%)-we call it global south countries in this study compared to high-income countries (67%)-non-global south countries is a challenge for public health. Since containing the spread of the coronavirus required global collaboration and access to vaccine to all people and countries, this study investigates the main contributing factors that determine global south countries to lag behind the non-global south countries in the uptake of the COVID-19 vaccines. The results of the study give ideas on actions that need to be taken by governments, policymakers, and pharmaceutical companies on the factors that contribute to the low uptake rate of the COVID-19 vaccine in low-income countries for ongoing and future pandemics.

## 3. Literature related to COVID-19 vaccines

The WHO report in 2022 shows that there are more than 20 vaccines essential to prevent life-threatening diseases since the outbreak of COVID-19 in 2019 (WHO 2022c). It has significantly reduced childhood mortality and improved the health, social and economic benefits of a community (Andre et al. 2008). Estimates in public health indicate that vaccination reduces 4 to 5 million deaths per year, and it is one of the most cost-effective measures (Rémy et al. 2015, WHO 2022c). Vaccinations are also essential for the economic growth of a country. The annual return of vaccinations on investment is 12% to 18% and is essential to reduce health inequality (Andre et al. 2008).

Experience of vaccination in developing countries have also shown a reduction in morbidity and mortality, empowerment, and economic benefits for women and children (Andre et al. 2008). For example, a population-based study conducted in Ukraine to assess the contribution of vaccination to the reduction of infectious mortality showed that vaccinations reduce 84.9% of tetanus-related mortality during 1944 to 2015 (Mokhort et al. 2018). Studies conducted in Bangladesh (Koenig et al. 1990) and Guinea-Bissau (Aaby et al. 1984) also showed a significant reduction in childhood mortality and improved short and long-term survival of children due to measles vaccinations. The mortality rate for measle-vaccinated children was 46% less compared to unvaccinated children (Koenig et al. 1990).

COVID-19 pandemic caused significant morbidity and mortality globally (Moghadas et al. 2021), and has created strong pressure on economic, social, and national health critical care systems in most countries. Particularly, developing countries are facing strong challenges because of vulnerable poor socioeconomic conditions and healthcare systems (Almeida 2020; WHO 2014; Cutler 2020; Emanuel et al. 2020). The lack of an adequate number of healthcare workers, laboratory test kits, and shortage of personal protective equipment (PPE) are the main challenges in responding to COVID-19 in most developing countries (WHO 2020). Moreover, there were tight travel and movement restrictions in several countries globally (Gupta et al., 2021), which have resulted in social, economic, and health consequences (Ahmed et al. 2020). Similarly, the COVID-19 pandemic significantly altered the health care seeking behavior of individuals especially for nonurgent and non-life-threatening conditions (Zhang, 2021).

Some other studies indicate that factors such as vaccine supply constraints, limited fiscal space for a bailout for access to vaccines, vaccine hesitancy, lower reported cases, and lack of governments effective approach to implement a range of strategies to increase access to vaccines in low-and-middle incomes countries contributed the low uptake rate of the COVID-19 vaccines in the global south countries.

The WHO Strategic Advisory Group of Experts (SAGE) on Immunization defines vaccine hesitancy as a delay in acceptance or refusal of vaccination despite the availability of vaccination services (Dror et al. 2020, MacDonald 2015, WHO 2019) also argue that while vaccine compliance remains variable and inconsistent, evidence from a diverse set of factors as well as different cases demonstrates the challenges and opportunities for public health. Others indicate factors that contribute to vaccine hesitancy including their coincidental temporal relationships to adverse health outcomes, unfamiliarity with VPD, and the lack of trust in corporations and public health agencies (Dror et al. 2020). According to (Saint-Victor and Omer 2013), the tendency of compliance with other VPD such as polio and measles reduced over time. Hence, even though vaccine hesitancy is one of the top threats to global health (WHO 2019), vaccine supply constraints are also a challenging factor in prolonging the virus and costing more lives.

### COVID-19 vaccine equity

The United Nations Development Program (UNDP) defines vaccine equity as “a means that vaccines should be allocated across all countries based on needs and regardless of their economic status”. It is equal access to vaccines without any distinction in race, religion, economic or social condition (UNDP et al. 2022). However, the COVID-19 pandemic has disproportionally affected racial and ethnic minorities (White and Grimm 2022; Scott et al. 2021).

Although several organizations have signed a vaccine equity declaration, the inequitable distribution of COVID-19 vaccines is still a major public health problem, especially in LMICs (WHO 2022b). To minimize disparities related to the COVID-19 vaccine, the promotion of vaccine equity and increasing access is essential (Scott et al. 2021).

The main reasons for vaccine inequities are vaccine hesitancy and vaccine access (White and Grimm 2022). Additional reasons for vaccine inequity include affordability of vaccines for poor countries, control of the vaccines by wealthy countries, control of global vaccine production by a small number of countries, and challenges with the distribution and administration of vaccines due to poor infrastructure in low-and middle-income countries (O’Leary and Tsui 2021). Moreover, an interrupted cold chain due to power supply problems and lack of adequate freezers is another major problem for getting COVID-19 vaccines to low-and middle-income countries (LMIC) (The Lancet Infectious 2021).

A report by UNDP, WHO, and the University of Oxford showed that low-income countries started vaccination an average of two months later than high-income countries. Although WHO calls for vaccine equity, there are still problems related to vaccine distribution to LMIC (WHO 2022g). For example, as of May 11, 2022, 72.08% (3 in 4) of people in high-income countries have been vaccinated with at least one dose. However, in low-income countries, only 17.4% (1 in 6) of people have been vaccinated with at least one dose (UNDP et al. 2022), although there is a higher willingness to take COVID-19 vaccines among people in LMIC (Solís Arce et al. 2021).

The inequity that exists in vaccine distribution has a profound impact on the socioeconomic status of countries, particularly in low-and middle-income countries (LMICs). This has exposed people living in LMIC to a high level of new COVID-19 cases (UNDP et al. 2022). Additionally, vaccine inequity could trigger the emergence of new COVID-19 variants (Afolabi et al. 2021; UNDP et al. 2022). Furthermore, a study conducted in Ohio, USA, showed the effectiveness of vaccine lottery program in increasing COVID-19 vaccination (Brehm et al., 2022)

### Covid 19 vaccine availability and distribution

The majority of vaccines administered so far are in high and upper-middle-income countries (WHO 2022b). The global unequal distribution of vaccines contributes to the low uptake of vaccine technology. For instance, as of May 11, 2022, the percentage of persons fully vaccinated with the last dose of primary series in high-income countries was 73.67% and it was 13.03% in lower-income countries (WHO 2022f).

As of May 11, 2022, there have been a total of 516,476,402 confirmed cases of COVID-19, including 6,258,023 deaths globally (Johns Hopkins University and Medicine, 2022). COVID-19 vaccinations are essential in mitigating new cases and deaths. A study conducted in the United States on the impact of vaccination on COVID-19 outbreaks showed a reduction in the overall attack rate from 9% to 4.6% and a reduction in adverse outcomes including death (Moghadas et al. 2021).

## 4. Materials and Methods

### Data Source and Setting

We extracted data related to the national COVID-19 vaccination coverage and uptake, COVID-19 new and cumulative cases and deaths, and vaccination policy for all countries from the Our World in Data, an online resource bringing together a large number of data sources from WHO’s individual national reports (Ritchie et al. 2020). Additional data sources include the WHO website for COVID-19 data (WHO 2022e), the COVID-19 Dashboard by the Center for Systems Science and Engineering (CSSE) at Johns Hopkins University (JHU) (CSSE JHU 2022), and Bloomberg graphics data on COVID-19 vaccine tracker (Bloomberg 2022). Furthermore, data on the first date of vaccination for each country was obtained from CNN Health (Shveda et al. 2022). The last date of data extraction was May 12, 2022.

Extracted data from the different sources were exported to STATA software (version 17) for analysis. The list of countries was categorized as global south and non-global south countries using information from United Nations (UN 2022). Accordingly, data from 149 global south and 59 non-global south countries were included in this study.

### Descriptive Analysis

Descriptive statistics like the frequency and percentage were used to present the different outcomes of the study. In addition, using country-level data, an event study analysis was used to examine the effects of an event (vaccination rates) on different outcomes of the study. Accordingly, the main event for this study is the vaccination against COVID-19. The occurrence of this event was considered as day 0. Tables and figures were created considering the outcomes of the study. For continuous variables, data were presented using descriptive statistics such as mean, and standard deviation for normal distribution, and median and interquartile range (IQR) for not normally distributed. For categorical variables, frequency and percentage were used. Normality tests were checked using visual inspection and the Shapiro-Wilk and Kolmogorov-Smirnov statistical tests. For outcomes that violated the normality test, analysis techniques including median and inter-quartile range (IQR).

### Estimation Strategy

Using country-level data, an event study analysis was used to examine the effects of an event (COVID-19 vaccination rollout) on outcomes like new COVID-19 cases and new COVID-19 deaths between 1 January 2020 to 12 May 2022. This method utilizes the variation in the treatment timing across different countries at different periods. Accordingly, the main event for this study is the start of vaccination against COVID-19 for each country. Similar to a difference-in-differences design, the event study analysis allowed use to estimate the effects in the context of a natural experiment, comparing the pre-post initial rollout changes in COVID-19 outcomes in countries with earlier rollouts versus changes in these outcomes in countries what initiated their vaccination rollout later. In relation to the COVID-19 research, several studies have also applied an event study method. Verma et al. (2021), for instance, used the event study method to analyze the impact of COVID-19 on different sectors of the economy in India using lockdown announcement day by the government as an event. Similarly, Isynuwardhana and Putri (2021) evaluate whether there are significant differences in abnormal returns, trading volume activity, and bid-ask spread before and after the announcement of COVID-19 in Indonesia by applying an event study method. Thus, applying an event study method is robust for our analysis to examine the effects of an event (COVID-19 vaccination rollout) on new cases and deaths related to COVID-19 (outcomes).

We estimated the effects of the initial vaccination rollout on the count of daily new COVID-19 cases or new COVID-19 deaths, controlling for total cases, number of ICU patients, number of hospital patients, number of cardiovascular death rate, median age, number of people aged 65 or older, number of people aged 70 or older, and diabetes prevalence, new tests, number of female smokers, and number of male smokers, country fixed effects, and day fixed effects. The regression equation is as follows.

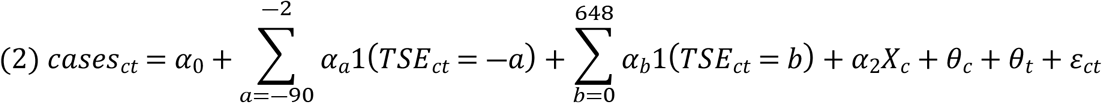

As a standard event study analysis, the model includes a set of event times (TSE) for country s, indicating the number of days relative to the initial COVID-19 vaccination rollout of country s (the event of interest). The reference period for estimating the initial rollout effects was 1 day before the initial vaccination rollout. Country fixed effects were used to adjust for time-invariant differences in countries’ capacity to contain the pandemic (e.g., workforce and acceptance toward new vaccines) and other geographic factors (Clarke and Tapia-Schythe 2021). The day fixed effects were used to control for global shocks such as outbreaks of new variants. The time event models were estimated by ordinary least squares regressions with country-clustered standard errors.

## 5. Results

### Vaccination policy

Based on the government policies on COVID-19 vaccination, we first group countries into six categories following (Ritchie et al., 2020). No availability (Category 0); Availability for ONE of following (Category1)-key workers/clinically vulnerable groups/elderly groups; Availability for TWO of following (Category2)-key workers/clinically vulnerable groups/elderly groups; Availability for ALL of following (Category3)-key workers/clinically vulnerable groups/elderly groups; Availability for all three plus partial additional availability (select broad groups/ages) (Categor4), and Universal availability (Category 5).

As of May 12, 2022 (the cut-off period for the current research), the majority (93.05%) of countries had a universal availability of vaccination policy. Whereas 4.81% of countries have availability for key workers/clinically vulnerable groups/elderly groups plus partial additional availability for selected broad groups or ages. When disaggregated by continents, all countries in Europe have universal availability of vaccination policy. In Asian and African countries, 91.5% and 82.4%, respectively, have universal vaccine availability. From global south countries, 9 (7.26%) countries had category 4 and 112 (90.32%) countries had universal availability policy. Overall, only 82.4% (42) African countries and 91.5% (43) Asian countries implemented universal availability policies, while 100% non-global South countries implemented this policy.

### COVID-19 Vaccination Distribution and Equity

The study also identified the variation between the global south and non-global south country categories in the number of total vaccinations, full vaccination, and total boosters. Although developing countries represent more than two-thirds (85%) of the global population (Schellekens and Sourrouille 2020), vaccination-related indicators are higher in developed than in developing countries.

As of May 2022, the median time in weeks since the first dose of COVID-19 vaccination varies across the two groups. It was 64 weeks for global south countries and 71.7 weeks for non-global south countries. As of May 2022, the median number of people who were fully vaccinated per hundred was 50.13 and 68.8 for global south and non-global south countries respectively. Additionally, the total number of boosters administered per hundred people in global south countries was 13.02, and it was 45.7 in non-global south countries (Table 1).

**Table 1.**
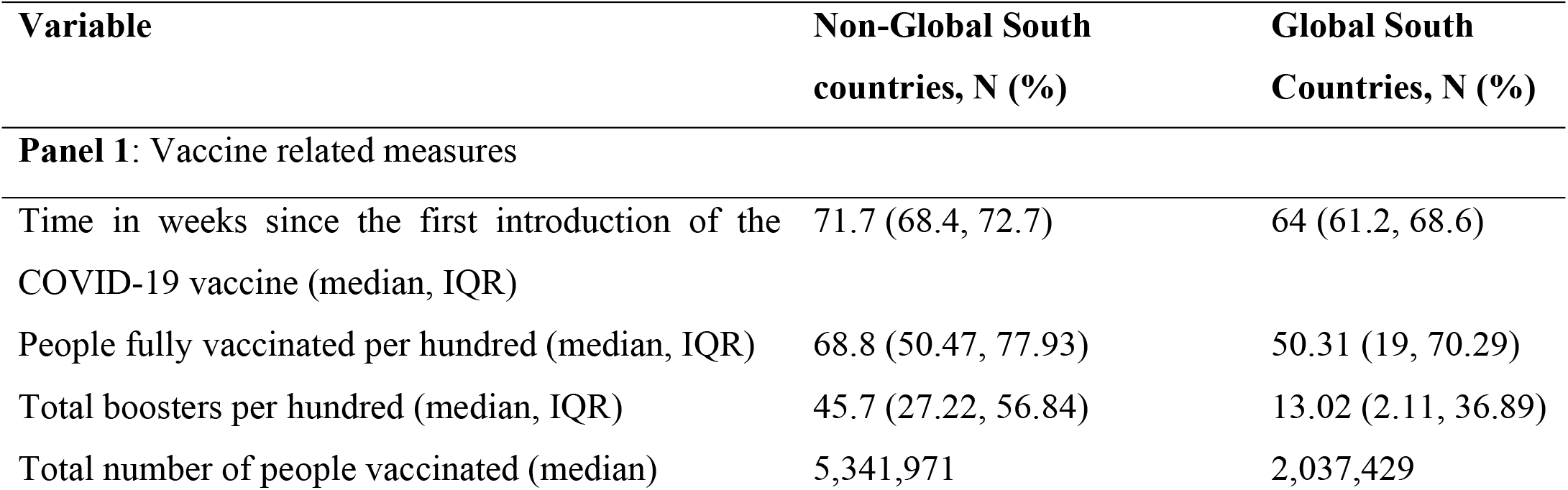
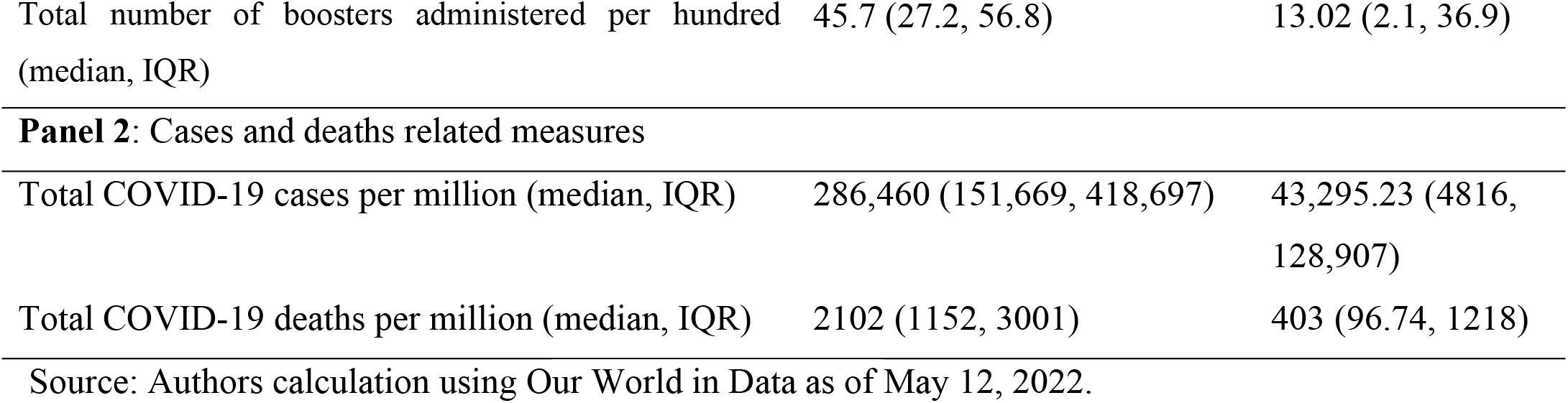
Distribution of time since first COVID-19 vaccination, people full vaccinations per hundred, total boosters, total COVID-19 cases, and deaths for global south and non-global south countries.

| Variable | Non-Global South countries, N (%) | Global South Countries, N (%) |
| --- | --- | --- |
| **Panel 1: Vaccine related measures** |  |  |
| Time in weeks since the first introduction of the COVID-19 vaccine (median, IQR) | 71.7 (68.4, 72.7) | 64 (61.2, 68.6) |
| People fully vaccinated per hundred (median, IQR) | 68.8 (50.47, 77.93) | 50.31 (19, 70.29) |
| Total boosters per hundred (median, IQR) | 45.7 (27.22, 56.84) | 13.02 (2.11, 36.89) |
| Total number of people vaccinated (median) | 5,341,971 | 2,037,429 |
| Total number of boosters administered per hundred (median, IQR) | 45.7 (27.2, 56.8) | 13.02 (2.1, 36.9) |
| **Panel 2: Cases and deaths related measures** |  |  |
| Total COVID-19 cases per million (median, IQR) | 286,460 (151,669, 418,697) | 43,295.23 (4816, 128,907) |
| Total COVID-19 deaths per million (median, IQR) | 2102 (1152, 3001) | 403 (96.74, 1218) |

Source: Authors calculation using Our World in Data as of May 12, 2022.

The mean number of total vaccinations per hundred people was lower in global south countries compared to non-global south countries. Similarly, the mean number of people fully vaccinated per hundred and mean of total boosters administered per hundred were lower in global south countries compared to non-global south countries (Table 1).

### COVID-19 vaccination and outcomes

The treatment timing for the various countries included in this study is presented in the table below. More than two-third (76.27%) of non-global south countries and only 35.14% of global south countries started COVID-19 vaccination in the first 6 months of vaccination start period. Whereas a large percentage of global south countries (62.84%) vs 22.03% of non-global south countries started during 7-12 months after the start of global vaccination (Table 2).

**Table 2.** Distribution of countries by COVID-19 vaccination start period since first global vaccination for non-global south and global south countries.

| Vaccination start period | Non-Global South countries, N (%) | Global South Countries, N (%) |
| --- | --- | --- |
| 0-6 months | 45 (76.27) | 52 (35.14) |
| 7-12 months | 13 (22.03) | 93 (62.84) |
| 13-17 months | 1 (1.69) | 3 (2.03) |

Source: Authors calculation using Our World in Data as of May 12, 2022.

We also estimated the change in the different outcomes of the study before and after the introduction of COVID-19 vaccinations for global south countries. Although overlapping 95% confidence intervals in the before and after the COVID-19 vaccination period, there was a higher number of cases per million after COVID-19 vaccination compared to the period before the COVID-19 vaccination. Although not statistically significant, the median number of new cases per million and the new ICU patients per million people before and after the COVID-19 vaccination increased after the start of vaccination (Table 3).

**Table 3.** Distribution of outcomes before and after the COVID-19 vaccination in global south countries.

| **Variables** | **Before COVID-19**<br>**vaccination** | **After COVID-19**<br>**vaccination** |
| --- | --- | --- |
| New cases per million people (median, IQR) | 1.51 (0, 19.07) | 7.13 (0, 87.1) |
| New ICU patients per million (median, IQR) | 11.03 (0.95, 46.74) | 15.5 (4.64, 44.21) |
| Weekly ICU admission (median, IQR) | 316 (264, 558) | 274.5 (134, 743) |
| Life expectancy in years (mean, SD) | 70.64, 7.48 | 71.31, 7.37 |
| New cases per million (median, IQR) | 1.51 (0, 19.1) | 7.13 (0, 87.1) |
| New deaths per million (mean, SD) | 0.66 (2.46) | 1.42 (5.94) |

Source: Authors calculation using Our World in Data as of May 12, 2022.

### Event study plots

The event study plots below show the impact of COVID-19 vaccination on the number of new cases and new deaths. Accordingly, there was a statistically significant reduction of COVID-19 new cases after the first COVID-19 vaccination rollout compared to the baseline in global south countries (before starting the COVID-19 vaccination), p-value <0.001 (Figure 1).

**Figure 1.**
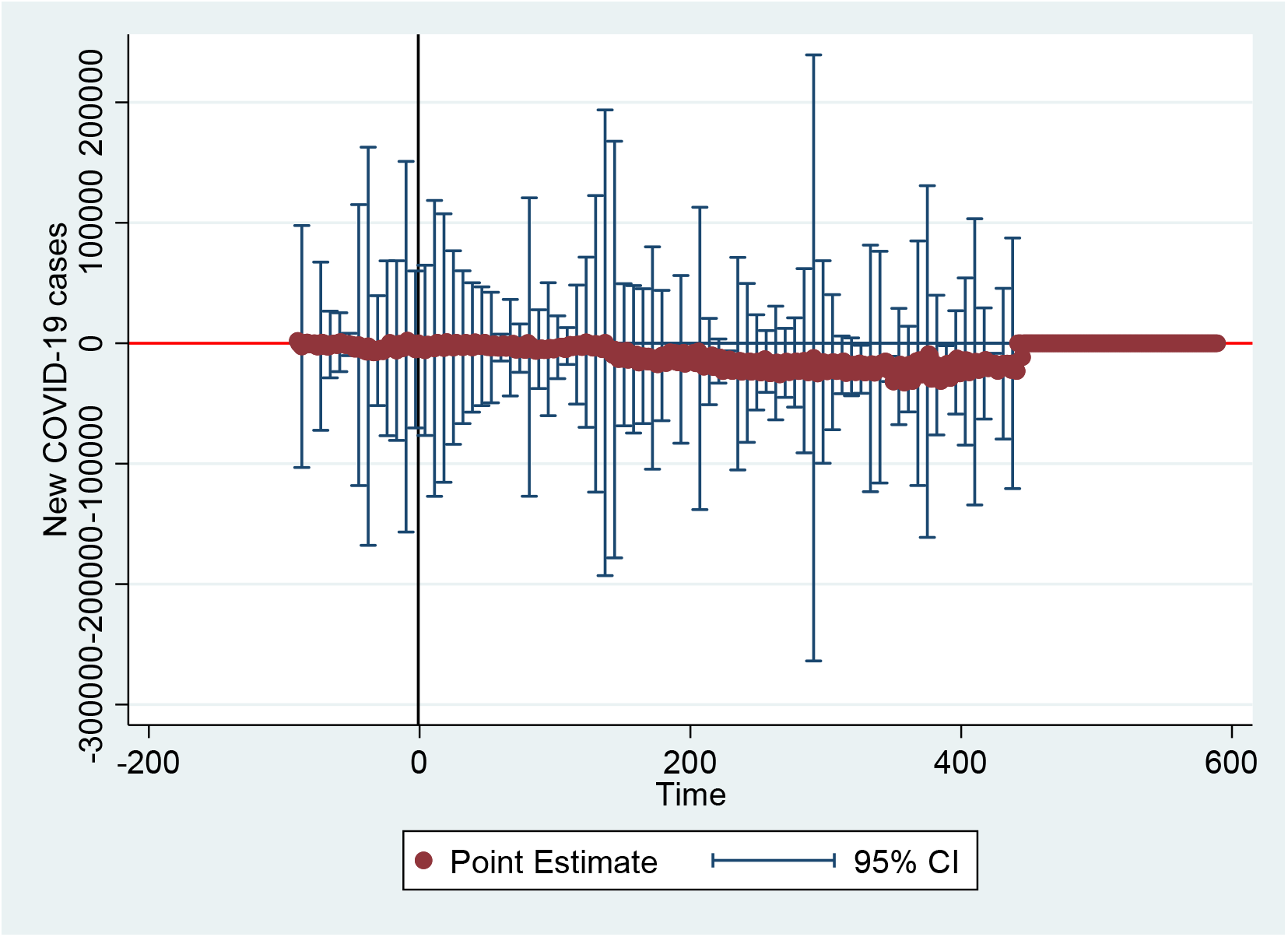
Event study plot associated to DD model of new COVID-19 cases in global south countries (Time is in days).

The event study analysis for new COVID-19 related deaths as an outcome also adjusted for new COVID-19 cases and the factors indicated above. As presented in the figure below, there was a statistically significant reduction in the number of new COVID-19 deaths after the COVID-19 vaccination period compared to the number before the vaccination period in global south countries, p-value <0.001 (Figure 2). The summary statistics of the event study plot for global south countries is presented in Table 4.

**Table 4.** Summary statistics of the event study plot associated to DD model of new COVID-19 cases and deaths in global south countries

| Variables* | Covid-19 new cases outcome |  |  | Covid-19 new deaths outcome |  |  |
| --- | --- | --- | --- | --- | --- | --- |
|  | Coefficient | Robust SE | P-value | Coefficient | Robust SE | P-value |
| Total cases | .0052333 | 5.02e-10 | <0.001 | -9.50e-06 | 6.59e-12 | <0.001 |
| ICU patients | -13.32753 | 5.79e-07 | <0.001 |  |  |  |
| Cardiovascular death rate | -15.7996 | 8.95e-06 | <0.001 | .4991023 | 7.23e-08 | <0.001 |
| Constant | -3260.379 | .0019766 | <0.001 | -162.7227 | .000031 | <0.001 |

Source: Our World in Data as of May 12, 2022.
\*Variables like female smokers, male smokers, median age, aged 65, aged 70 older, and diabetes prevalence were omitted from the new cases and deaths analysis because of collinearity.

**Figure 2.**
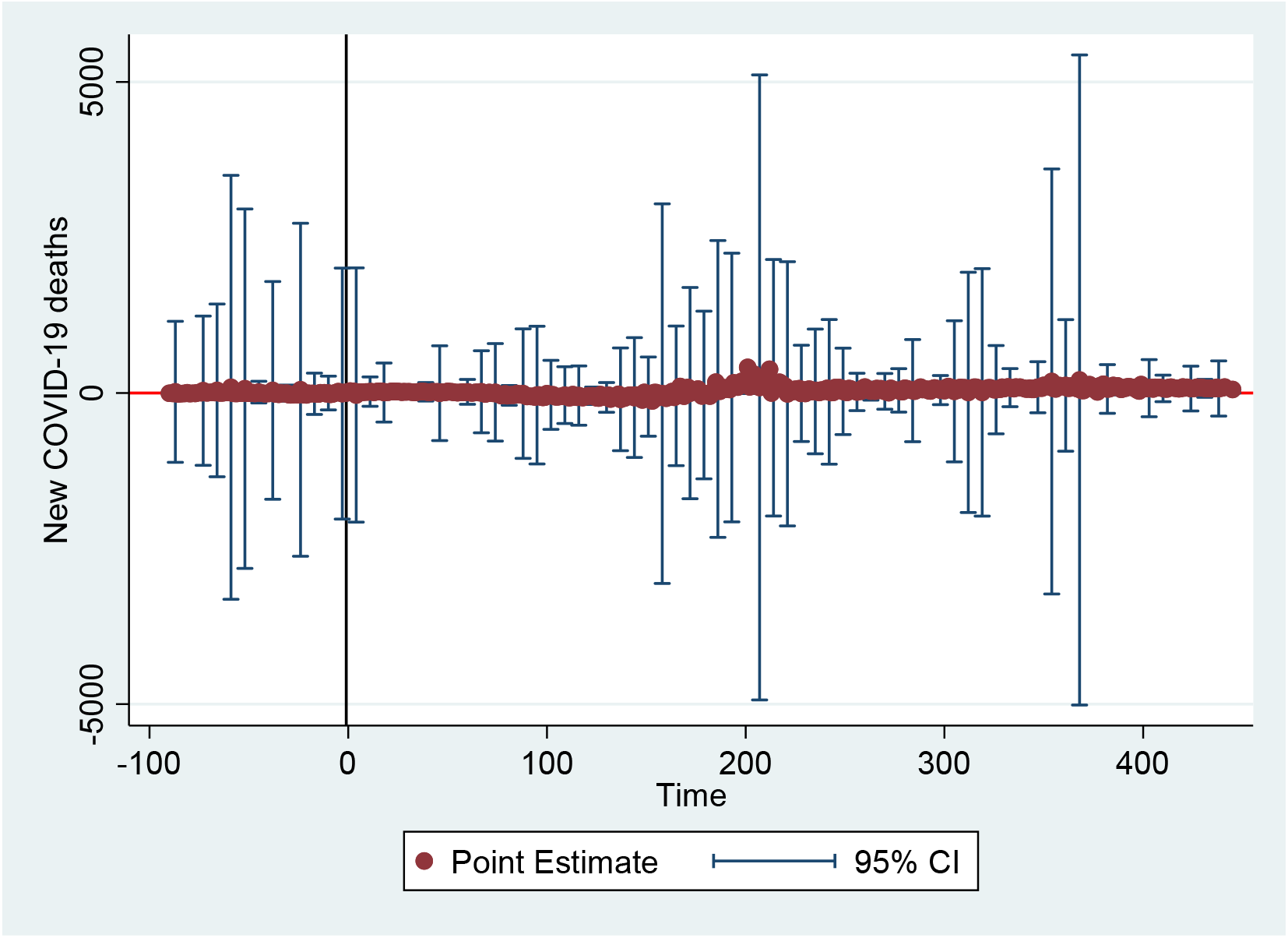
Event study plot associated to DD model of new COVID-19 deaths in global south countries (Time is described in days).

For non-global south countries, although reduction in the number of COVID-19 new cases after the vaccination rollout it was not statistically significant. Similar finding was also found for the COVID-19 related deaths (Figure 3 & 4). The summary statistics of the event study plot for non-global south countries is presented in Table 5.

**Table 5.**
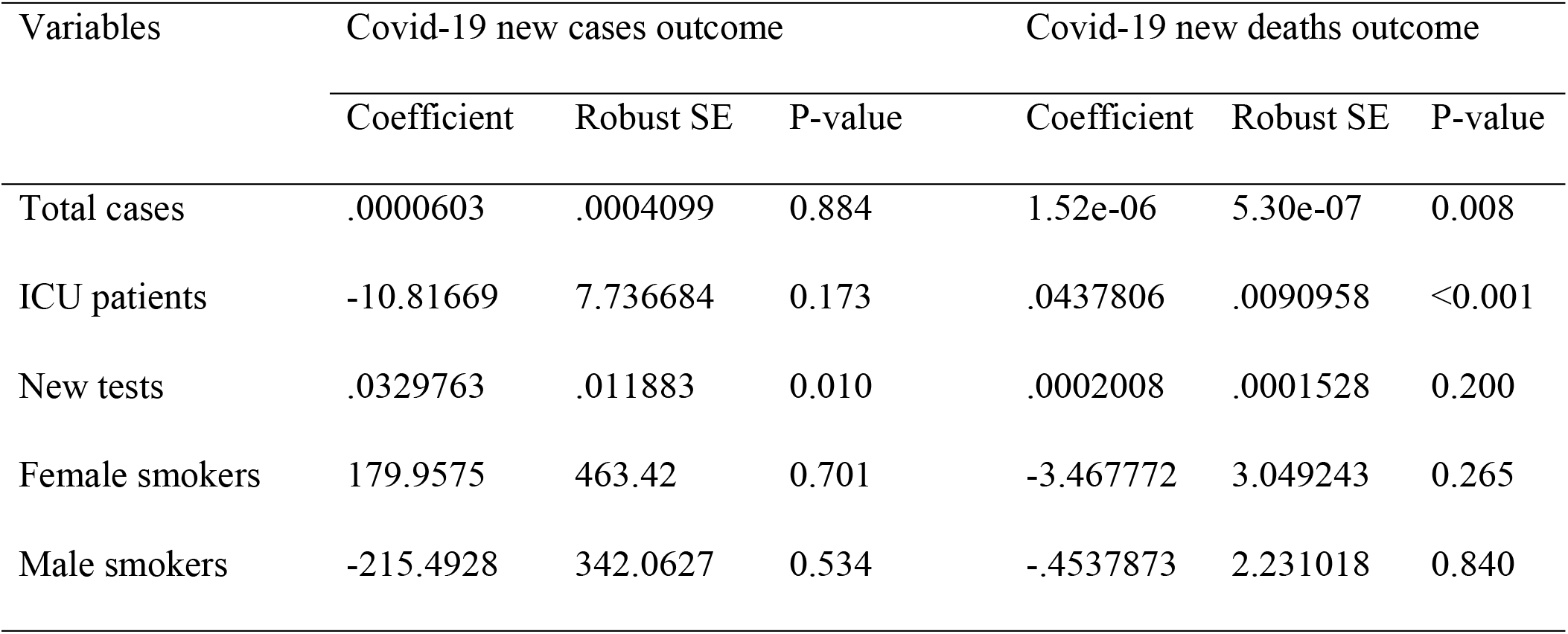
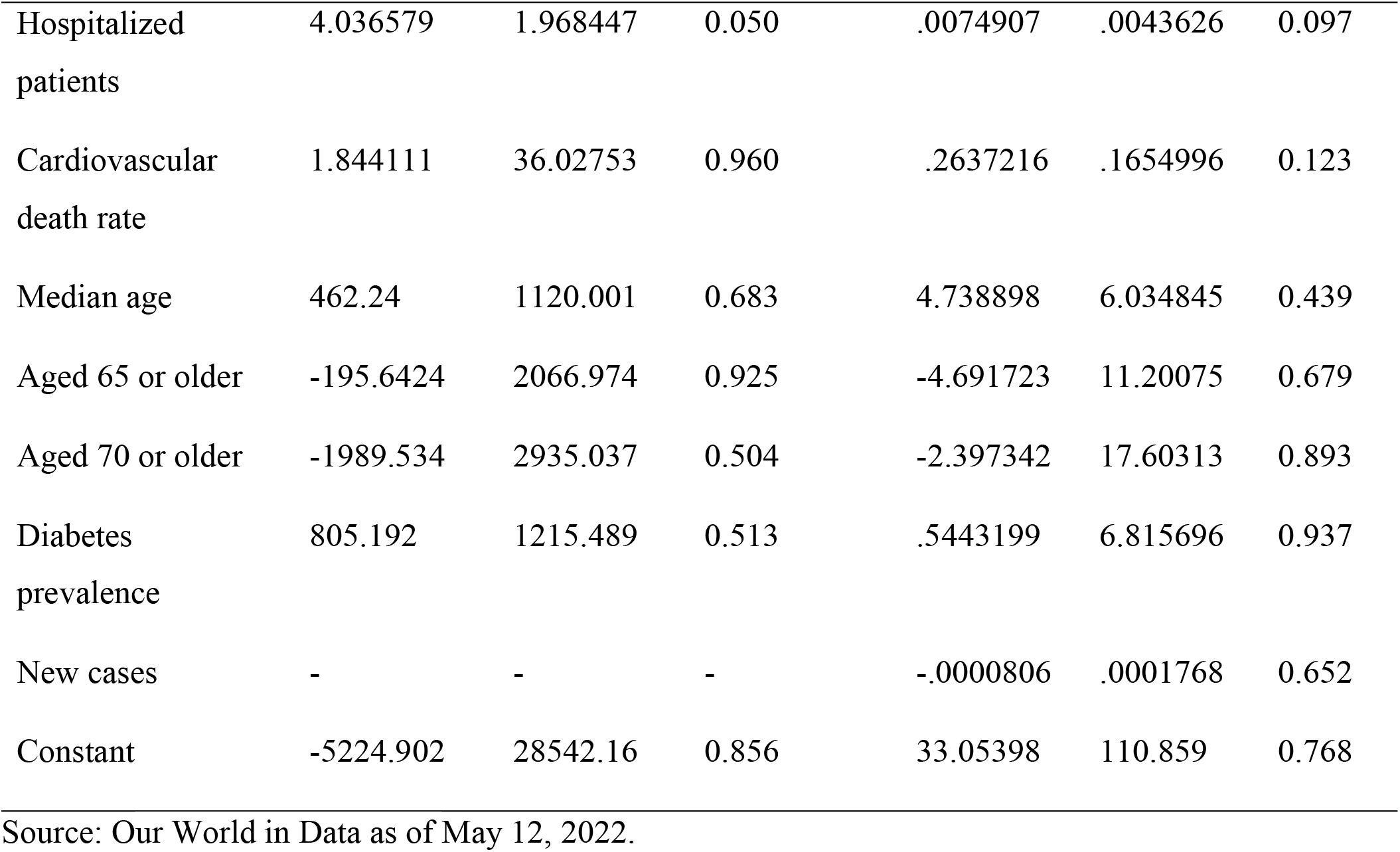
Summary statistics of the event study plot associated to DD model of new COVID-19 cases and deaths in non-global south countries

| Variables | Covid-19 new cases outcome |  |  | Covid-19 new deaths outcome |  |  |
| --- | --- | --- | --- | --- | --- | --- |
|  | Coefficient | Robust SE | P-value | Coefficient | Robust SE | P-value |
| Total cases | .0000603 | .0004099 | 0.884 | 1.52e-06 | 5.30e-07 | 0.008 |
| ICU patients | -10.81669 | 7.736684 | 0.173 | .0437806 | .0090958 | <0.001 |
| New tests | .0329763 | .011883 | 0.010 | .0002008 | .0001528 | 0.200 |
| Female smokers | 179.9575 | 463.42 | 0.701 | -3.467772 | 3.049243 | 0.265 |
| Male smokers | -215.4928 | 342.0627 | 0.534 | -.4537873 | 2.231018 | 0.840 |
| Hospitalized patients | 4.036579 | 1.968447 | 0.050 | .0074907 | .0043626 | 0.097 |
| Cardiovascular death rate | 1.844111 | 36.02753 | 0.960 | .2637216 | .1654996 | 0.123 |
| Median age | 462.24 | 1120.001 | 0.683 | 4.738898 | 6.034845 | 0.439 |
| Aged 65 or older | -195.6424 | 2066.974 | 0.925 | -4.691723 | 11.20075 | 0.679 |
| Aged 70 or older | -1989.534 | 2935.037 | 0.504 | -2.397342 | 17.60313 | 0.893 |
| Diabetes prevalence | 805.192 | 1215.489 | 0.513 | .5443199 | 6.815696 | 0.937 |
| New cases | - | - | - | -.0000806 | .0001768 | 0.652 |
| Constant | -5224.902 | 28542.16 | 0.856 | 33.05398 | 110.859 | 0.768 |

Source: Our World in Data as of May 12, 2022.

**Figure 3.**
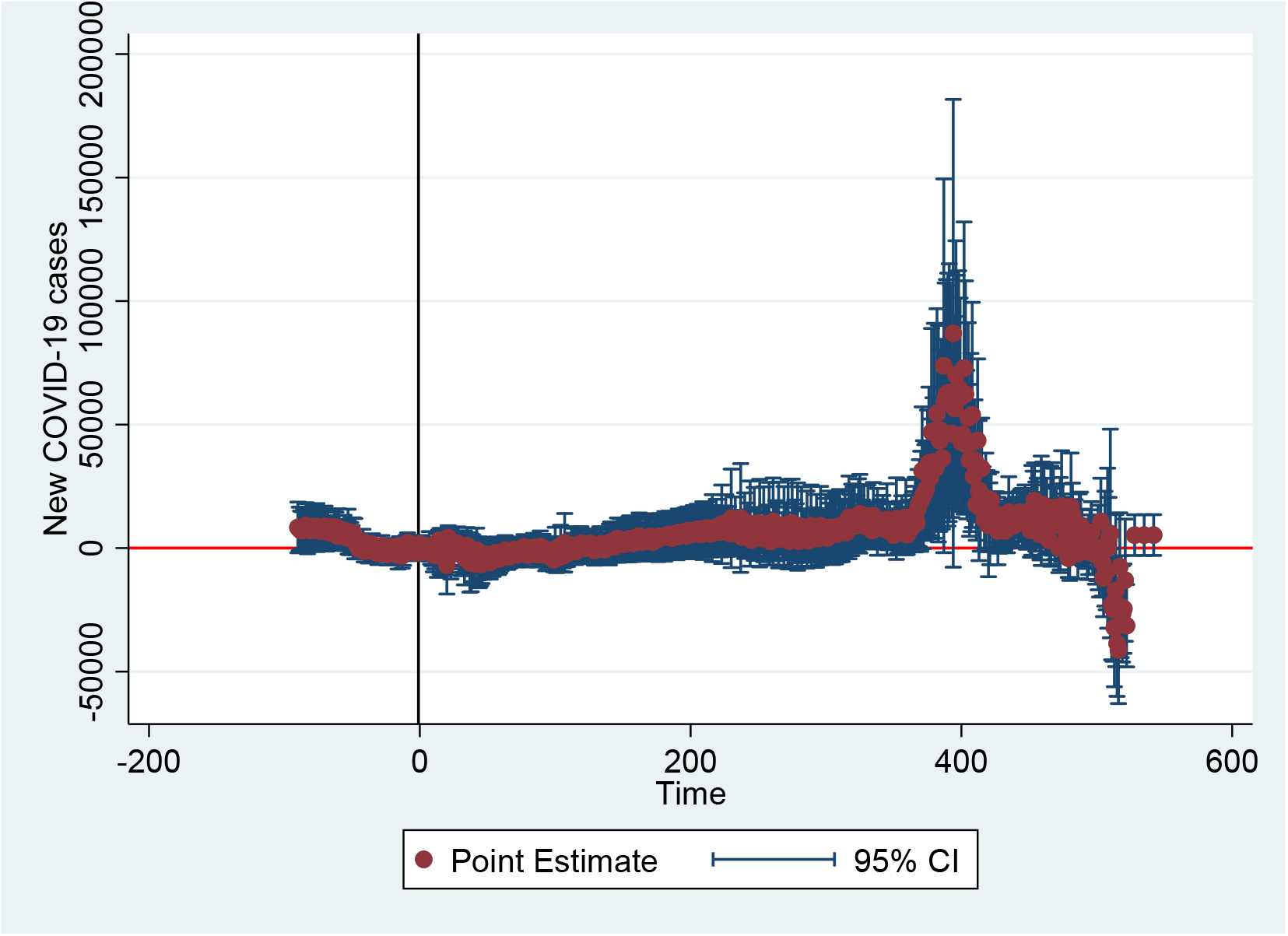
Event study plot associated to DD model of new COVID-19 cases in non-global south countries (Time is in days).

**Figure 4.**
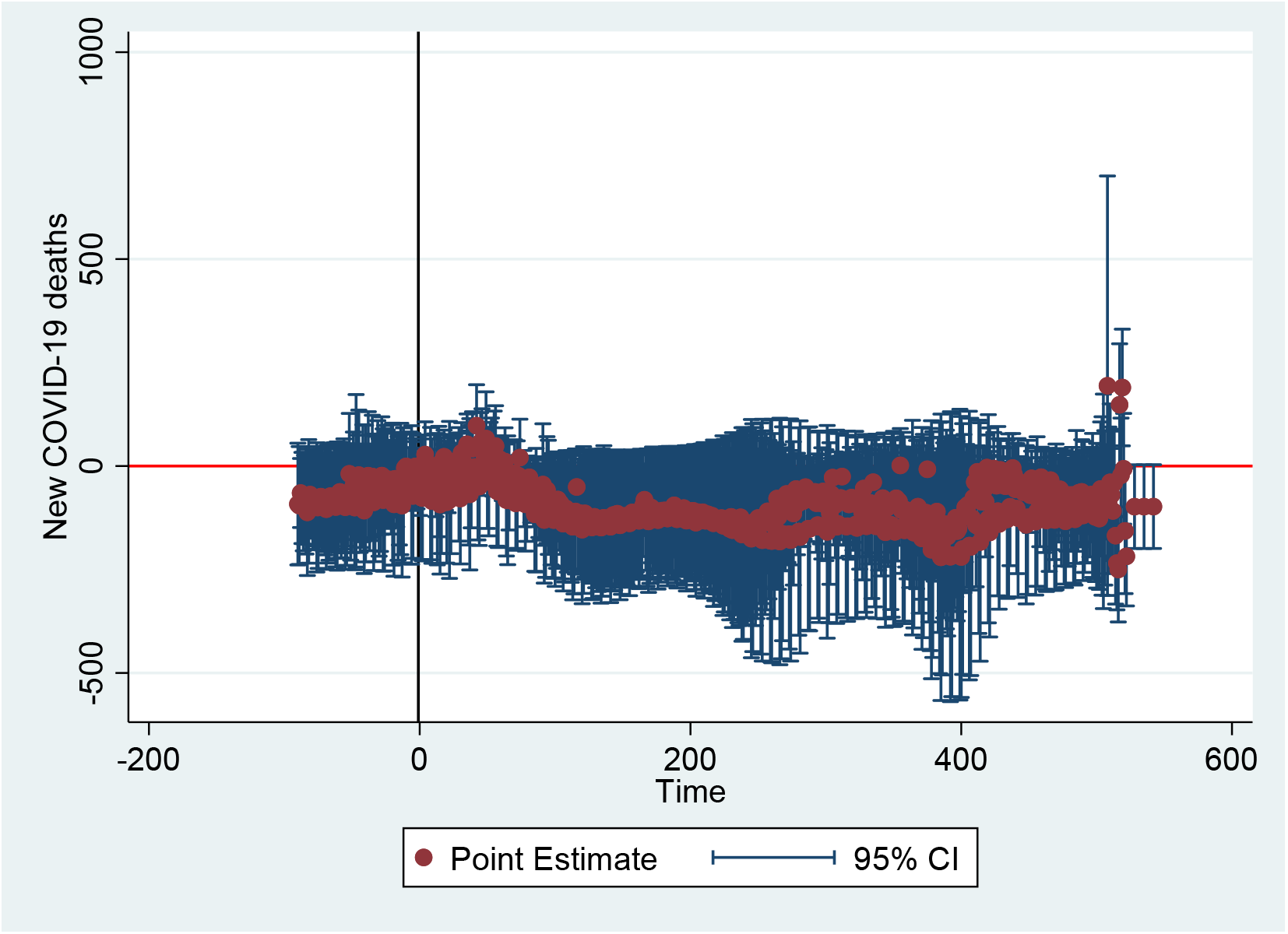
Event study plot associated to DD model of new COVID-19 deaths in non-global south countries (Time is in days).

## 6. Discussions

This study was conducted to track the COVID-19 vaccine equity, distribution, and cases in global south countries using country-level data from Our World in Data using an event study analysis. Our study finding showed issues related to vaccine inequity between global south and non-global south countries. There was variation in the proportion of countries with universal availability of vaccines in both regions, higher in non-global south countries. Similarly, non-global south countries introduce COVID-19 vaccines early than global south countries. The event study analysis also identified a significant reduction of COVID-19 new cases and deaths in global south countries after the first vaccination rollout compared to the baseline.

Most countries campaign COVID-19 vaccination to control the disease burden, including the new cases and deaths. As the result of these campaigns and uptake of the vaccinations, there have been reduction in the rate of COVID-19 transmission rate. Similar to the findings of our study, a study conducted in the US also showed a reduction in the COVID-19 deaths and economic benefits as the result of the vaccination campaigns (Gupta et al. 2021).

The current study found that all countries in non-global south countries and 90.32% of global south countries had universal availability of COVID-19 vaccines. Similarly, the mean number of fully vaccinated people per hundred and mean total boosters administered were lower in global than non-global south countries. This could be related to the accessibility of vaccines to high-income countries and significant delays in vaccine access in LMICs (Ramachandran et al. 2021). Therefore, it is essential to address the coverage gap in the COVID-19 vaccinations in global-south countries to better control the pandemic.

Our event study analysis also showed a significant reduction of COVID-19 new cases after the first COVID-19 vaccination rollout compared to the period before starting the COVID-19 vaccinations in global south countries. This supports the strong efficacy and effectiveness of COVID-19 vaccines in reducing morbidity and mortality as reported in previous clinical trial (Rubin and Longo 2020). Similar findings were also reported in a previous study (Gupta et al. 2021).

When comparing the reported number of COVID-19 cases per million before and after the first COVID-19 vaccine rollout in global south countries, our study showed that there was higher number of cases in the period after the start of vaccination. This could be because of an improved reporting system and testing in the period after the start of COVID-19 vaccination compared to the period before it. In poor countries, especially during the period after the start of the pandemic, there was an under-reporting of cases and deaths due to the lack of testing and reporting mechanisms (OXFAM International 2022).

The current study also showed that the median number of new cases per million and the new ICU patients per million people increased after the start of vaccination. This could be because of the high number of total COVID-19 cases during the same period due to the second wave of COVID-19. Additional factors for this could also include infection prevention policies of countries, human behavior, and change in the coronavirus because of mutations and variants (Maragakis 2021). However, the median number of weekly ICU admissions reduced from 316 before the COVID-19 vaccination to 274.5 after the COVID-19 vaccination period (Table 3). A similar study also showed a significant reduction in mortality (by 54%) and ICU admission (23.3%) among people who received at least one dose of the COVID-19 vaccine (Selvavinayagam et al. 2022).

Our study has several strengths. Our study used different data sources to estimate the change in the COVID-19 new cases and deaths before and after the vaccine introduction. We also adjusted the event-study analysis by different characteristics that could affect the cases and deaths.

The study is also not without limitations. Although our study tried to adjust for different factors that could affect the COVID-cases and deaths, additional factors could also contribute the variation in the coverage and effectiveness of the vaccination administration. Factors like the socioeconomic status of the population, availability of the infrastructures may have also contributed to the variation.

## 7. Conclusions

This study was conducted to track the COVID-19 vaccine equity and distribution in global south countries. An event study analysis was also conducted to examine the effects of COVID-19 vaccination on new cases and deaths related to COVID-19. The findings showed that there was variation in the timing of COVID-19 vaccine introduction in the global south and non-global south countries. The median time since the introduction of the first COVID-19 vaccine in global south countries was almost eight weeks later than non-global south countries, 64 weeks for global south countries, and 71.7 weeks for non-global south countries. A comparison of outcomes before and after the first COVID-19 vaccination rollout also showed a reduction in the number of ICU weekly admissions and almost one-year increase in the mean of life expectancy in global south countries, although not a significant change.

The event study plots also showed a statistically significant reduction in the number of new COVID-19 cases and deaths in global south countries. There is unequal distribution of COVID-19 vaccinations between non-global south and global south countries. There are significant variations in the number of vaccinations, full vaccination, and booster doses administered between the two groups of countries. Such inequality in vaccine distribution has prolonged the pandemic globally. Therefore, it is essential to improve vaccine access and distribution to global south countries to effectively control new COVID-19 infections and deaths.

## Data Availability

Availability of data and material: The datasets generated during this study are publicly available at the Our World in Data, Coronavirus Resource Center, repository https://ourworldindata.org/coronavirus

## Acknowledgments

First, we deeply thank the Johns Hopkins Coronavirus Resource Center for collecting the country-level data and make available for research use. We extend our gratitude to the Our World in Data team at the University of Oxford who contributed their time, energy and expertise, and publicize the data online (https://ourworldindata.org/coronavirus). We are also grateful to the Center for Effective Global Action (CEGA) at the University of California Berkey for providing us the opportunity to present this paper at the 2022 Africa Evidence Summit in Kigali, Rwanda, and receive valuable comments.

## Notes

**Declaration of Conflicting Interests:** The authors declared no conflicts of interest with respect to the research, authorship, and/or publication of this article.

**Funding:** The authors received no financial support for the research, authorship, and/or publication of this article.

### Competing Interest Statement

The authors have declared no competing interest.

### Funding Statement

The author(s) received no specific funding for this work.

### Author Declarations

This study used secondary data and did not require IRB approval.

